# Intense and Mild Wave of COVID-19 in The Gambia: a Cohort Analysis

**DOI:** 10.1101/2020.12.10.20238576

**Authors:** Anna Roca, MRCG/GG COVID-19 working group

## Abstract

**Background:** The SARS-CoV-2 pandemic is evolving differently in Africa compared to other regions, with lower transmission and milder clinical presentation. Reasons for this are not fully understood. Recent data from Eastern and Southern Africa suggest that transmission may be higher than measured. Detailed epidemiological data in different African settings is urgently needed.

**Methods:** We calculated cumulative rates of SAR-CoV-2 infections per 1,000 people at risk in The Gambia (2.42 million individuals) using publicly available data. We evaluated these rates in a cohort of 1,366 employees working at the MRC Unit The Gambia @LSHTM (MRCG) where systematic surveillance of symptomatic cases and contact tracing was implemented. Cumulative rates among the Gambian population were stratified by age groups and, among MRCG staff, by occupational exposure risk. SARS-CoV-2 testing was conducted on oropharyngeal/nasopharyngeal samples with consistent sampling and laboratory procedures across cohorts.

**Findings:** By September 2020, 3,579 cases of SARS-CoV-2 and 115 deaths had been identified; with 67% of cases detected in August. Among them, 191 cases were MRCG staff; all of them were asymptomatic/mild, with no deaths. The cumulative incidence rate for SARS-CoV-2 infection among MRCG staff (excluding those with occupational exposure risk) was 129 per 1,000, at least 20-fold higher than the estimations based on diagnosed cases in the adult Gambian population.

**Interpretation:** Our findings are consistent with recent African sero-prevalence studies reporting high community transmission of SAR-CoV-2. Enhanced community surveillance is essential to further understand and predict the future trajectory of the pandemic in Africa.

## INTRODUCTION

The SARS-CoV-2 pandemic has spread to six continents (1), with more than 45 million cases and 1.1 million deaths registered by end of October 2020. Even though Africa accounts for 15.6% of the worldwide population(2), it contributes only 3.9% cases (1.76 million) and 3.6% deaths (42,233) to the worldwide burden(1). Data suggest that, compared to the rest of the world, the pandemic is evolving differently in sub-Saharan Africa; the outbreak started later than elsewhere, probably due to the low intensity of international air traffic (3), and rapid closure of the international borders following the first few cases.

Importantly, COVID-19 severe disease seems to occur less frequently in Africa than in the rest of the world (4). Several factors have been proposed to explain this. Age is likely to be a major factor as older individuals are at higher risk of severe disease. Africa has an extremely young population, with a median age of 20 years compared to 43 years in Europe, 39 years in North America or 31 years in South America (5). However, variation of severity with age alone cannot completely explain the observed differences (4). Underreporting of both clinical cases and deaths due to limited systematic surveillance and of lack of systematic death registration may underestimate the true burden (4). Nevertheless, even if this may be true, local health systems, whose capacities to deal with COVID-19 patients are generally lower than in high resourced settings, did not become overwhelmed, even at the peak of the epidemic (6). Although potential avoidance of medical care during the pandemic as described in other regions (7) may partly explain the low number of hospitalized patients, the lower severity reported appears to be genuine and several biological and environmental factors have been proposed as potential contributing factors, i.e. lower prevalence of vitamin D deficiency, higher prevalence of helminths infection, higher coverage of BCG vaccination or cross-protection after infection by human coronavirus (8-10).

Recent sero-surveys conducted in Kenya, Malawi and South Africa showed that community transmission was several times higher than detected by surveillance, as 5% to 40% of the population had IgG SAR-CoV-2 antibodies (11-13). Such results highlight the need of robust epidemiological studies assessing the extent of community transmission in different African regions.

The Gambia is the smallest country in continental mainland Africa and is surrounded by Senegal, except for its narrow Atlantic coast. Although the first imported case was identified in The Gambia on March 17^th^, 2020, by June 30^th^ only 48 additional cases had been detected. Nevertheless, July saw a rapid increase of cases, with 3,579 cases by the end of September (1). The trajectory of the epidemic in The Gambia is different from that in Senegal (with a population about 7 times that of The Gambia) where community transmission was first reported in early April and almost 7,000 cases had been recorded by end of June (1). The MRC Unit The Gambia at the London School of Hygiene and Tropical Medicine (MRCG) is a biomedical research institution with 1,336 employees, mostly Gambians(14). Systematic surveillance and testing of MRCG staff with influenza-like symptoms together with contact tracing has been implemented during the pandemic, with the first case identified on July 18^th^. We have considered MRC staff as a cohort able to provide additional insights into the nature of the COVID-19 epidemic in The Gambia.

## METHODS

### The Gambia

The Gambia has an estimated population of 2.42 million in 2020, with a median age of 17.8 years. Around 41.9% of the population are between 20 and 64 years-of-age. About 95% of the population is Muslim and illiteracy rate is high across the country. Around 59% of the population live in urban and peri-urban settings, mainly at the coast (Figure 1). The climate is typical of the sub-Sahel region, with a long dry season from November to May and a short rainy season between June and October. Maximum temperature is high throughout the year (between 30^°^C and 34^°^C; lowest during the rainy season) while minimum temperature increases during the rainy season (16^°^C −20^°^C during the dry season and 22^°^C −24^°^C during the rainy season) (15). Humidity can surpass 80% during the rainy months (16).

**Figure 1.**
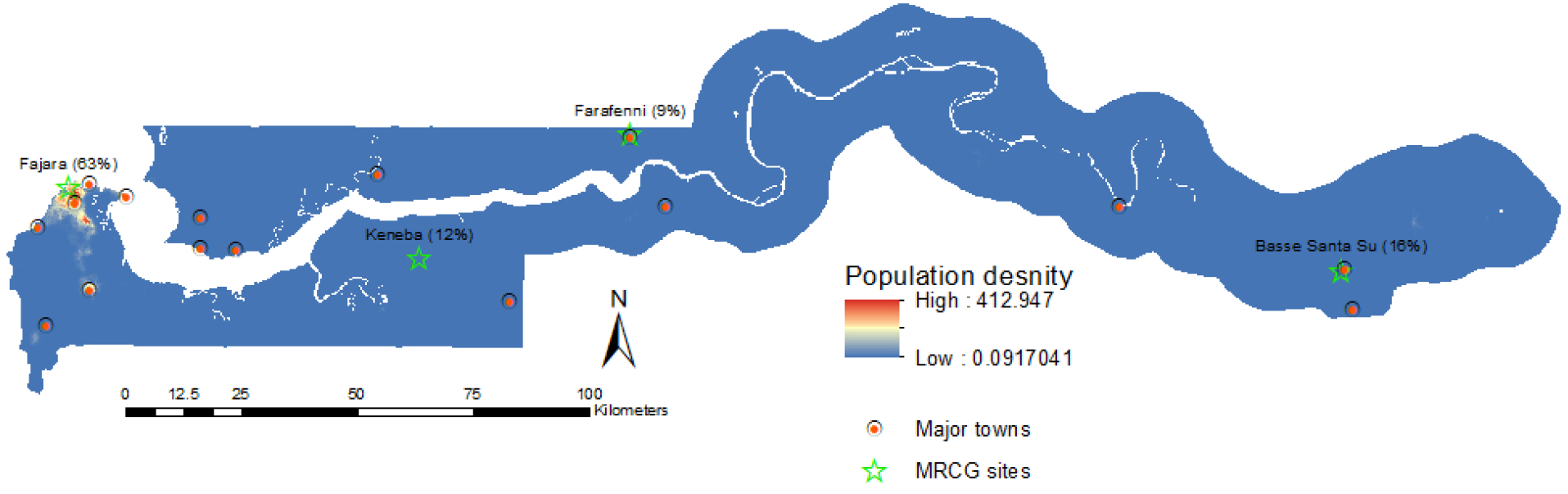
Gambian map of population density and including the MRCG research sites distributed across the country.

The government of The Gambia is the main health provider and health care delivery has three tiers, based on the primary health care strategy. There are four tertiary hospitals, 38 health centers at the secondary level and 492 health posts at the primary level. The system is complemented by 34 private and nongovernmental organization (NGO) clinics.

### COVID response in The Gambia

Shortly after the first case, the country closed its international land, sea and air borders (19^th^ March) and soon after (27^th^ March) a state of emergency was declared, with closure of schools, non-essential shops, places of worship and many working places. Initial SAR-CoV-2 testing by PCR was focused on the identification of imported cases and the tracing and isolation of their contacts, especially among travelers from Senegal as the long borders are permeable with people often living between the two countries. As the epidemic progressed, testing facilities were established at strategic locations in the most densely populated parts of the country, mainly in the western urban areas, and individuals were encouraged to present for testing if symptomatic or following known contact. All identified cases were isolated in designated facilities regardless of symptoms until considered non-infectious as per WHO guidelines. Contacts were traced and quarantined in hotels during the early part of the outbreak (April – July), after which self-isolation at home was allowed.

### MRCG Unit

MRCG is a biomedical research institution which also provides outpatient and inpatient clinical care to the local population through its clinical services department (CSD). The Unit had 1,336 employees as of August 2020, distributed between the coast (mainly in Fajara, n=845), Keneba (n=158), Central River Division (mainly Farafenni, n=116) and Upper River Division (mainly Basse, n=217) (Figure 1). Staff work in different environments, including those that are field-based (drivers, fieldworkers, nurses, research clinicians [n=715, 53.5%]), office-based (administrative, operations, data-management, statistics [n=334, 25.0%]), laboratory-based (n=177, 13.2%). A small number of the MRCG staff provides health care to the general population attending the CSD (n=110, 8.2% of the total MRCG staff).

CSD is one of the two Gambian hospital facilities able to hospitalize severe COVID-19 patients (with 42 beds for COVID-19 patients), both for MRCG staff and the general population. From the start of the epidemic, all staff were trained to wear appropriate personal protective equipment (PPE) based on international guidelines.

MRCG staff underwent a clinician-administered risk assessment and those deemed to be at high or medium risk were advised to work from home and were excluded from high risk clinical areas.

### Surveillance and contact tracing among MRCG staff

Since July, MRCG established processes for testing all symptomatic staff (including their families and contacts) and staff known to have been exposed to confirmed cases. A staff hotline was set up, manned by doctors, for easy access to this service and for answering any questions or concerns about symptoms and how to access services. Contacts were called to confirm exposure and then tested 3-5 days after the last exposure. Regardless of a negative test results, all exposed staff were quarantined for 14 days with staff testing positive isolated in line with WHO recommendations.

Besides the testing described above, CSD staff were offered weekly PCR-based testing irrespective of symptoms

### Sample collection

Nasopharyngeal (NPS) and/or oropharyngeal (OPS) samples were collected with swabs (FLOQSwabs, COPAN Diagnostics), and placed in single tubes containing universal transport medium (UTM: COPAN Diagnostics). Samples were delivered to the laboratory within 24 hours. Sampling methods were comparable across cohorts with similar operational procedures and training.

### Laboratory methods for COVID detection

MRCG laboratories collaborated with the Gambian National Public Health Laboratories (NPHL) to support national testing throughout the country during the epidemic, with the same laboratory methods and assays.

In anticipation of the spread of the outbreak to the West African subregion, MRCG attended a CDC-Africa regional training workshop on diagnosis of 2019 novel coronavirus held in February 2020 in Dakar, Senegal. Thereafter, laboratory protocols for processing and testing suspected coronavirus-infected samples using the WHO guidelines were established in The Gambia (17;18). The same procedures and assays were transferred to the NPHL.

The standard test for COVID-19 diagnosis in The Gambia is the real-time RT-PCR of SARS-CoV-2 specific viral gene sequences. In the early stages of the outbreak, RT-PCR diagnosis was based on the Berlin Charité Laboratory protocol (19) which targets the RNA-dependent RNA polymerase (RdRP) and Envelope protein (E) gene. Subsequently, tests kits, primarily the DAAN and SANSURE kits, were donated to the NPHL; both target the open reading frame (ORF-1ab) and the nucleocapsid gene (N) coding regions.

Sample inactivation and downstream RNA extraction was done with commercially available kits following the manufacturers’ protocol. Initial extractions were manually done with the QIAamp Viral RNA Mini Kit (QIAGEN) or the IndiSpin Pathogen Kit (INDICAL BIOSCIENCE). When donations to the public health system became available, the DAAN and SANSURE kits, (DAAN Gene Co., Ltd of Sun Yat-sen University; Sansure Biotech Inc Hunan, China) were included. As the outbreak progressed and daily sample numbers increased, automated RNA extraction system on the QIAcube HT (QIAGEN) was implemented. In all cases, 200µL of UTM sample was processed and the RNA eluted in 50µL-80µ depending on the extraction kit. RT-PCR analysis was conducted with 5µL of extracted RNA in 25µL of reaction mix containing, reaction buffer, one-step reverse transcriptase enzyme, either the Takara One Step PrimeScript III RT-PCR Kit (TAKARA Bio) or SuperScript III Platinum™ One-Step qRT-PCR Kit (INVITROGEN, Thermo Fisher Scientific)” and the primer and probe mix.

Samples were defined as positive if amplification of any viral gene occurred by the 40^th^ cycle and with all the controls amplifying as appropriate.

### Definition of cases

A COVID-19 case was any individual with a positive RT-PCT of SARS-CoV-2 from a NPS/OPS sample irrespective of symptomatology.

### Statistical analysis

We calculated cumulative rates per 1,000 individuals at risk. Different rates for the Gambian population were calculated: (i) entire population, (ii) rates for the population 20-64 year and, (iii) rates for the population 20-64 years living in major cities and towns (for the latter all clinical cases aged 20-64 years were considered in the numerator but only the proportion living in major towns were considered in the denominator; place of residence for the SARS-CoV-2 cases is not publicly available). For MRCG, rates are reported stratified by occupational clinical exposure (those working at the CSD versus the rest). In addition to occupational clinical exposure, surveillance for CSD staff was more intense due to routine testing, irrespective of symptoms or known exposure.

### Databases

National Gambian data was extracted from the publicly available John Hopkins University COVID-19 database(20).

### Ethics

National Gambian data was extracted from the publicly available John Hopkins University COVID-19 database(20). The Gambian Government / MRCG Joint Ethics committee approved the study (Ref L2020.E37).

## RESULTS

### Baseline characteristics

Table 1 shows the main epidemiological characteristics of the Gambian population and the MRCG cohort. In the MRCG cohort, individuals aged >60 years are underrepresented while urban residents are overrepresented.

**Table 1.** Epidemiological and demographic characteristics of The Gambian population and MRCG staff.

| Baseline characteristics |  | The Gambia<br>N (%) | MRCG staff<br>N (%) |
| --- | --- | --- | --- |
| Age groups N(%) | <20 years | 1,338,834 (55.4%) | 2 (0.2%) |
|  | 20-60 years | 1,012,584 (41.9%) | 1,291 (98.5%) |
|  | >60 years | 65,250 (2.7%) | 17 (1.3%) |
| Age median (years) |  | 17.8 years | 37.5 years |
| Gender (N,% male) |  | 1,193,834 (49.4%) | 86 (55.1%) |
| Location (N % living in main towns/cities)* |  | 1,435,308 (59.4%) | 903 (67.6%) |

\* For the MRCG staff we have considered the working place rather than the living place.

### Samples tested and positivity rates

#### National Data

From the start of the epidemic until Sep 30^th^, 3,590 out of 17,885 samples tested positive for SARS-CoV-2. The positivity rate was low before July (1.6%; 40 out of 3,095) and high from July to September (23.7%; 3,499 out of 14,790) (19;21). Figure 2 shows the daily number of swabs collected in the country and the positivity rate in August and September 2020, when the number of samples and positivity rate were highest. During these two months, the number of daily swabs collected varied between 28 and 524 (median of 184). Positivity rate also varied substantially, between less than 5% and above 50%. About 67% of the confirmed cases were detected in August and, overall, 60% were aged less than 40 years(21).

**Figure 2.**
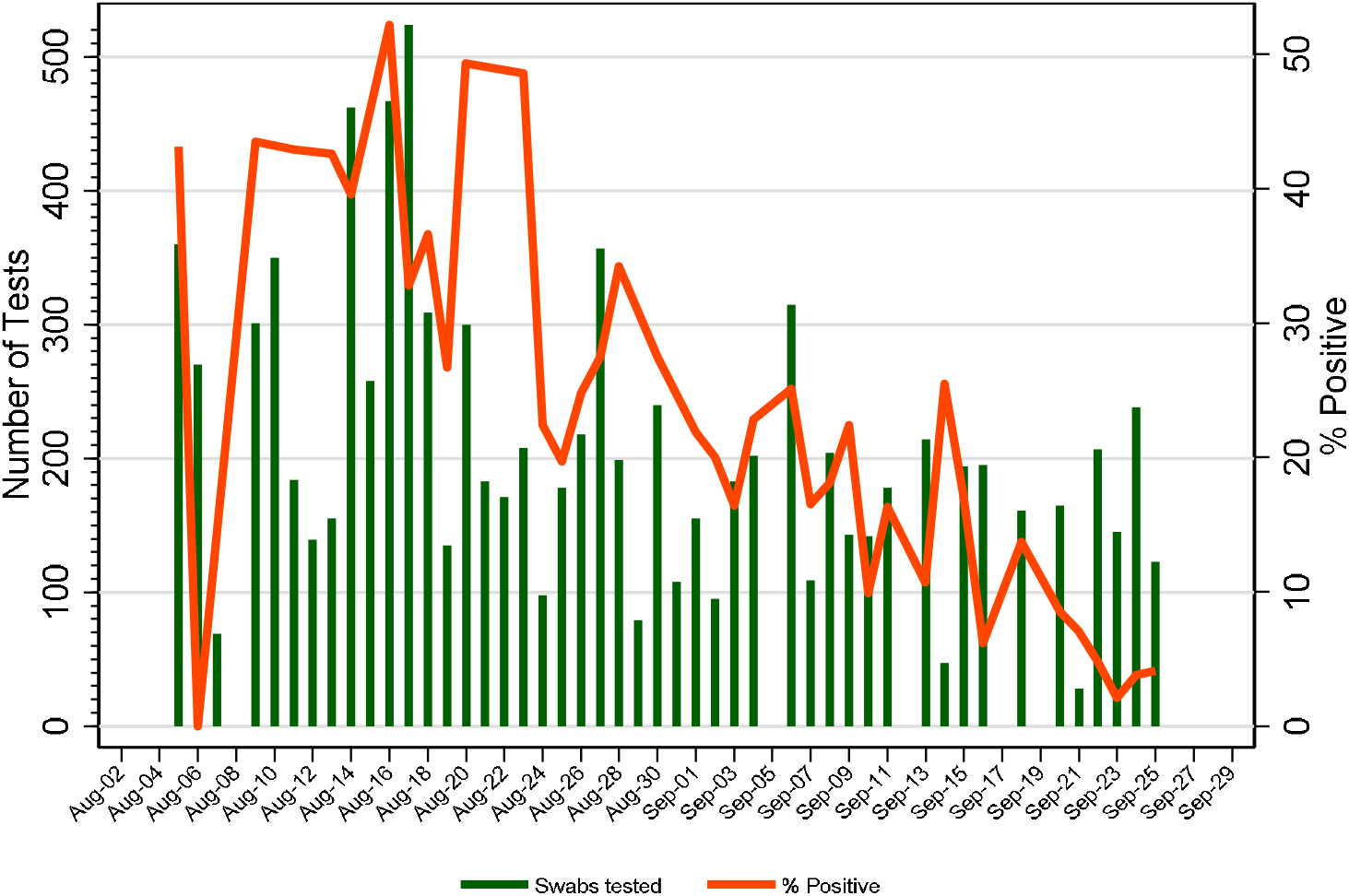
Number of daily COVID-19 tests in The Gambia and % of positive samples during the two months of intense transmission.

#### MRCG cohort

Between July and Sep 30^th^, 191 out of 937 samples collected tested positive for SARS-CoV-2 (20.4%). Sixty percent of the confirmed cases were detected in August; median age was 36 years.

### Rates of infection and deaths

#### National data

By the end of September, the cumulative rate in the Gambian population was approximately 1.5 per 1,000, slightly higher for adults 20-65 years - between 3 per 1,000 (overall) and 5 per 1,000 (when considered only adult population living in urban areas) (Figure 3a). During the same period, 115 COVID-19 deaths were recorded across the country.

**Figure 3.**
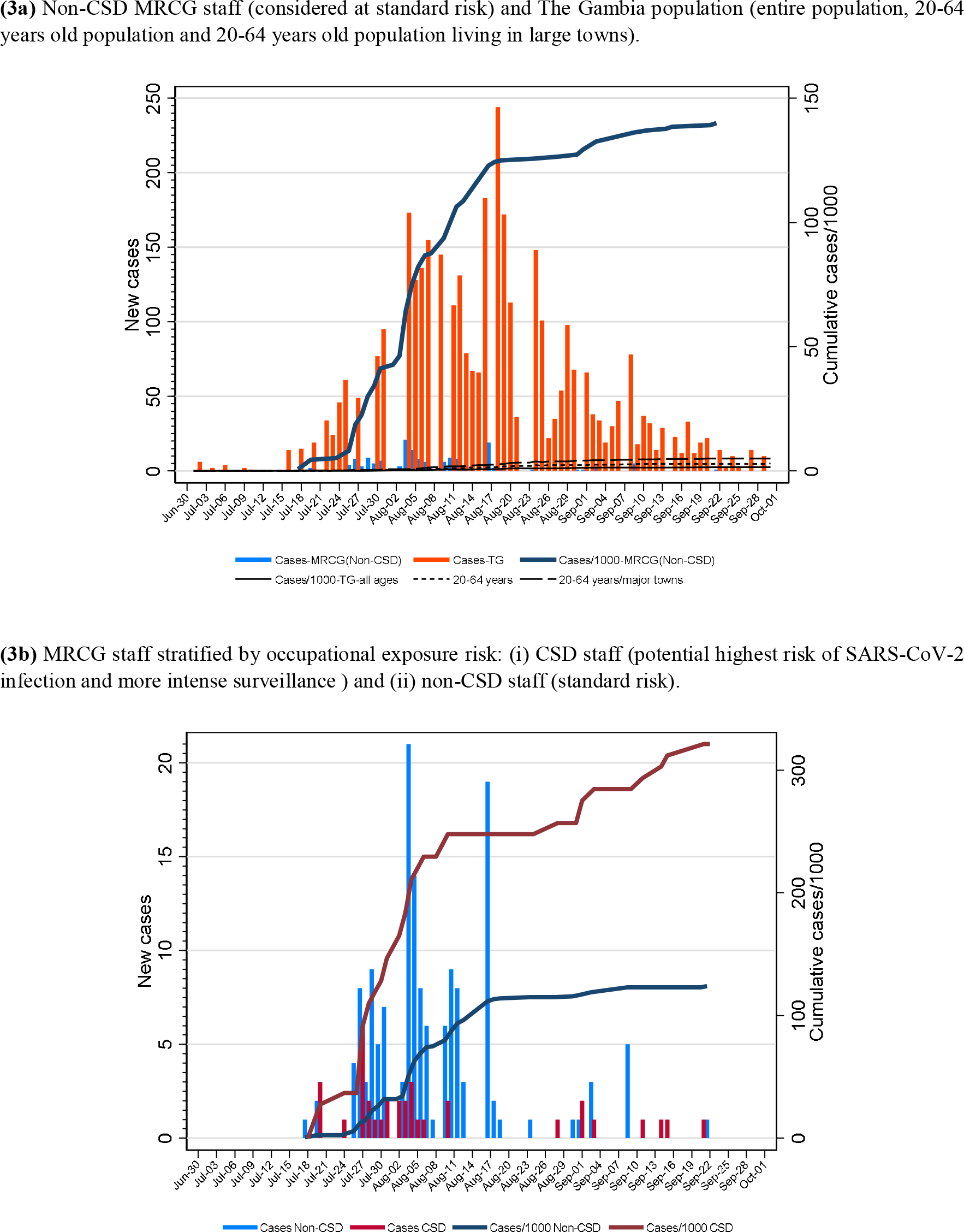
Daily cases and cumulative rate of infection per 1,000 until end of September 2020. TG=The Gambia; *<u>Note Y-axis for Figures 3a and 3b are different</u>*.

#### MRCG cohort

Stratified analysis shows that rates among CSD staff was 2.6 times higher than non CSD staff (considered representative of the general population risk) (Figure 3b). By the end of September, the cumulative risk for non-CSD MRCG staff was approximately 124 per 1,000 population (Figure 3b). All the 191 MRCG confirmed cases were either asymptomatic or with mild symptoms; with none meeting WHO criteria for moderate or severe pneumonia. There were no deaths.

## DISCUSSION

In The Gambia, the SARS-CoV-2 pandemic arrived later than in most countries in the world (in July), had a short and intense first wave (with 67% of cases occurring in August; with 1 out of 7 of individuals from our MRCG cohort being infected), and most cases were asymptomatic or mild. During this peak, positivity rate of SARS-CoV-2 for samples tested was very high at above 20%.

The late start of the epidemic is probably the result of the early closure of national borders, including for air travel, and of the identification and isolation of infected individuals who continued to enter the country from Senegal. This was complemented by contact tracing and by the provision by the provision of facilities for quarantine by the government. The relative impact of these measures, together with others implemented during the state of emergency, and of behavioral changes (social distancing, handwashing) is hard to quantify. Nonetheless, they seem to have been key in allowing the country to prepare its response and minimize potential harm. The sudden increase of cases coincided with the major Muslim feast of Eid-Ul Adha (locally called Tobaski on 30^th^ July in 2020), during which traveling and family gatherings were common, although the number of cases had already started to increase in July. Although climate in The Gambia is hot throughout the year, the peak epidemic coincided with the months of highest daily humidity and highest minimum temperature but lowest maximum temperature (15;16). Data on how temperature and humidity affects transmission are contradictory (22;23). In The Gambia, climate conditions may have had an indirect effect on transmission as during the rainy season people are more likely to spend time indoors. These are also the months with highest respiratory virus transmission (24).

Through the systematic testing of the MRCG staff cohort, including of asymptomatic contacts and mildly symptomatic cases, we are likely to have more robust estimates of the actual rates of SARS-CoV-2 infection in The Gambia than that available from the general population. The rate of SARS-CoV-2 in non-CSD MRCG staff (124 per 1,000 individuals) was more than 20-fold higher than that reported for the general population (between ∼1.5 and 5 per 1,000 individuals), even within the more intensely populated coastal area of The Gambia. Assuming that the Gambian urban adult population had similar exposures and transmission as our MRCG cohort, we would expect at least 75,000 infections in the 20-64 years age group living in main towns (out of the 601,394 individuals). This estimation contrasts sharply with the 3,579 cases reported during the same period across the country and in all age groups. Such discrepancy may be partly explained by the high occurrence of asymptomatic or mildly symptomatic infections, along with the national testing strategy based on passive case detection and of targeting symptomatic individuals. Indeed, such differences are consistent with recent seroprevalence studies from Eastern and Southern Africa that suggest higher rates of community infection compared to those estimated by passive case surveillance. In South Africa, three weeks after the COVID-19 peak, 40% of HIV-positive pregnant women had anti-SARS-CoV-2 antibodies (12). In Kenya, a retrospective survey of blood donor samples found that 1 in 20 adults had SARS CoV-2 antibodies between April and June 2020 (13). In Malawi, seroprevalence in a cohort of 500 health care workers sampled between May and June 2020 was 12.3%, concluding that with the observed prevalence the number of predicted deaths was eight times the number of reported deaths (11). In a smaller study in Nigeria (n=113), 45% of front line health care workers had SARS-CoV-2 antibodies (25). In The Gambia, more than 30% of the CSD staff became infected by 30^th^ September. Rates among CSD staff were higher than the rest of the MRCG cohort. This probably reflects a combination of factors: (i) stronger surveillance and (ii) occupational clinical exposure exacerbated by traveling to work. The weight of each factors is difficult to estimate. However, higher prevalence among health care workers has been reported in Europe (26).

The common mild disease is also reflected by the low occupancy of our hospital beds reserved for severe COVID-19 patients. However, this may also indicate avoidance of SARS-CoV-2 testing as a result of stigmatization (7) as observed in other regions. Indeed, 30% of tests of the 115 counted deaths were done post-mortem in patients hospitalized in non-COVID-19 health facilities. Without an official registration system for deaths, the overall toll of COVID-19 associated deaths is difficult to quantify and the real number may be several times higher.

Using MRCG staff as a cohort to estimate infection rates in the Gambian population has important limitations. Although cases in the general population and the MRCG cohort showed similar timelines and the size of the MRCG cohort is relatively large, the MRCG can be considered a cluster. The level of education and the monthly income of the staff is above that of the general population. MRCG staff live mainly in urban areas, where transmission tends to be higher (27) but also lives in less crowded environments, with better access to water and sanitation which would protect them from infection. In addition, MRCG developed policies and undertook significant levels of staff education related to COVID-19, reinforced messages related to social distancing, hand washing and the wearing of face masks at work as well as in the community. Finally, given the nature of the MRCG’s work, the level of background understanding on infectious diseases, even among staff not directly involved in research, is likely to be higher than in the general population. Moreover, the robust surveillance should have further limited transmission due to the rapid identification and isolation of cases. On the other hand, there was no moderate or severe COVID-19 case among the MRCG staff. This mild presentation was not modified by treatment as for instance, not a single member of the MRCG staff met WHO criteria for hospitalization and less so for oxygen supplementation or dexamethasone treatment. The prevalence of risk factors for severity should be similar between MRCG staff and the rest of the population except for the lower prevalence of individuals above 60 years of age, a main risk factor for severity and mortality.

The low (compared to other continents) levels of severe disease observed underlines the importance of minimizing the potential collateral damage of the COVID-19 pandemic in Africa. Such damage includes diversion of financial and personnel resources from other services to COVID-19 response, health seeking behaviour changes, reduced availability of medicines for acute and chronic diseases and disruptions of routine vaccination services (28-32). The pandemic has also worsened, particularly in low- and middle-income countries (33), the economic stability of households and lower food security; mitigating short and mid-term impact should be prioritized.

In conclusion, SARS-CoV-2 transmission in The Gambia has, to date, been intense over a short period of time. Reassuringly, the disease seems less severe than in high-income countries in Europe, North America and Asia. It is unclear whether a second wave of infection may occur given that the causes of the sudden increase of cases in July are not clear. We strongly encourage the continuous protection of health care workers with appropriate PPEs while strengthening surveillance systems around the country to promptly detect another sudden increase of cases. Country-wide sero-prevalent surveys are also necessary to understand micro-epidemiology of infection in different age groups and places. However, engaging with the community to mitigate collateral damage of the pandemic should take priority. It is also important to investigate the major drivers that shape the epidemic so differently in Africa than in some high-income regions as this understanding should help in modelling adequate interventions, both in low- and in high-income countries.

## Data Availability

The MRC Unit The Gambia at the LSHTM has a governance department were all enquiries regarding access to data need to be submitted.

## AUTHORS CONTRIBUTION

Study design: AR, UDA

Analysis plan: AR

First draft of the manuscript: AR, EC, UDA

Statistical analysis: AR, NM

Verification of Data: AR, NM

Figures: NM

Manuscript review: AB; AKS, AP, AR, AJ, AV, BAw, BAb, BK, BN,BS; CC, DN; EmO; EC, EO, EN, EU, FA, FO, HJ, HB, KB, KF, NH, NM, MA, MON, MJ, OA, OS, SJ, TsD, UO, WO, YO, BS, BM, CR; SS, ShJ, UDA, WO, YS, AS, CR, MB, MD.

Critical interpretation of the data: AP, AR, AJ, BK, BN, DN, EC, EU, HB, KF, MA, MMA, UDA, WO, AS, CR, MB.

Data collection: AB; AKS, AV, BAw, BAb, BS; CC; DN; EC, EmO; EO, EN, EU, FA, FO, HJ, HB, KB, KF, NH, MON, MJ, OA, OS, SJ, TsD, UO, WO, YO, BS, BM, CR; SS, ShJ, YS.

Approved final version: AB, AKS, AV, AR, AR, AJ, BAw, BAb, BK, BN, BS, CC, DN, EC, EU, EmO, EO, EN, FA, FO, HJ, HB, KB, KF, NH, NM, MON, MA, MMA, MJ, OA, OS, SJ, TdS, UO, UDA, WO, YO, AS, BS, BM, CR, MB, MD, SS, ShJ, YS

